# Do Electronic Nicotine Delivery Systems (ENDS) Help Smokers Quit? A Causal Bayesian Additive Regression Tree Analysis of PATH Study Waves 4 to 6

**DOI:** 10.64898/2026.11.23.26368804

**Authors:** Shu Xu, Jennifer Hill, Luchang Cui, Pan He, Yang Feng, Nan Jiang, Raymond S. Niaura

## Abstract

**Objective:** Observational evidence on the effects of ENDS use on cigarette smoking cessation remains mixed, partly because of potential confounding. This study aimed to estimate the effects of ENDS use on subsequent current smoking over a two-year follow-up.

**Methods:** We analyzed longitudinal data from Waves 4 to 6 of the Population Assessment of Tobacco and Health (PATH) Study (collected between 2016/12 and 2021/11), comprising a cohort of 4,792 US adults who were current established cigarette smokers at Wave 4. Using Bayesian Additive Regression Trees (BART) for causal inference, we estimated the effect of current ENDS use on subsequent current smoking, adjusting for a comprehensive set of 25 pre-exposure covariates. We characterized ENDS exposure as no use (0 days), some-day use (1 – 29 days), and daily use (30 days) in the past 30 days. Current cigarette smoking outcome was defined as past-30-day smoking, two years after the exposure measure.

**Results:** Over a two-year period, daily ENDS use substantially reduced the percentage of current smoking compared with no use (estimate = −15.7%, 95% CI: −21.5% to −9.9%) or compared with some-day use (estimate = −15.0%, 95% CI: −22.3% to −7.8%). In contrast, some-day ENDS reduced a much smaller percentage compared with no use (estimate = −0.6%, 95% CI: −3.2% to −2.0%). Sensitivity analyses indicated that the estimated effect of daily ENDS use was robust to moderate-to-large unmeasured confounding. There was no evidence that these effects varied meaningfully by sex, race/ethnicity, annual household income, sexual orientation, internalization, externalization, or age.

**Conclusions:** Among established cigarette smokers, daily ENDS use, compared with some-day or no use, reduced the probability of current smoking. These findings support the potential role of ENDS as a harm reduction tool to facilitate smoking cessation among adults.

## Does Electronic Nicotine Delivery Systems (ENDS) Use Help Smokers Quit?

Cigarette smoking remains the leading cause of preventable disease, disability, and death in the United States (Lushniak et al., 2014). Quitting cigarettes is of crucial clinical and public health importance, as smoking is associated with severe health risks, including morbidity, premature mortality, and economic burden. Despite the availability of evidence-based smoking cessation programs and pharmacotherapies (e.g., nicotine replacement therapy, varenicline, and bupropion), their uptake often remains low and long-term success rates are even lower (VanFrank et al., 2024), highlighting the need for alternative, accessible, and highly effective strategies to support individuals to quit smoking.

ENDS, commonly known as e-cigarettes, have the potential to facilitate smoking cessation. Unlike traditional cigarettes, which burn tobacco, ENDS heat an e-liquid to produce aerosol that delivers nicotine. Because they do not burn tobacco, ENDS substantially reduce exposure to many harmful constituents of cigarette smoke, including tar and carbon monoxide (Azzopardi et al., 2016; Lindson et al., 2026; Travis et al., 2023; Wang et al., 2022). Although ENDS are not risk-free and can deliver addictive nicotine, completely switching from combustible cigarettes to ENDS is generally considered substantially less harmful than continued cigarette smoking (Azzopardi et al., 2016; NASEM, 2018; Travis et al., 2023). Consequently, many adult smokers use ENDS as a potential means of reducing or quitting smoking (Erinoso et al., 2024). National survey data indicate that most adult ENDS users are current or former cigarette smokers, with ENDS use particularly prevalent among younger adults (Kramarow & Elgaddal, 2023). Reflecting their growing uptake, ENDS were among the most commonly used tobacco products among U.S. adults in 2024, with 7.0% reporting current use, second only to cigarettes (9.9%) (Agaku, 2026; Agaku et al., 2014).

Evidence from randomized clinical trials increasingly supports the effectiveness of ENDS for smoking cessation. The 2026 Cochrane living systematic review, which evaluated 80 studies representing about 30,000 adult smokers, found high-certainty evidence that ENDS result in significantly higher quit rates at six months compared to nicotine replacement therapies, ENDS without nicotine, no support provision, or behavioral support only (Lindson et al., 2026). This clinical efficacy was also recently demonstrated in a landmark study in Switzerland (Auer et al., 2024). UK’s National Institute for Health and Care Excellence recommended ENDS as an effective cessation option (NICE, 2025). U.S.-based physicians were also recommended to actively integrate ENDS into cigarette cessation practice (Leavens et al., 2026).

Evidence from observational studies on the effectiveness of ENDS for cessation remains mixed. A recent review of 28 articles encompassing 38 analyses from the Population Assessment of Tobacco and Health (PATH) Study found that 21 analyses reported associations consistent with a cessation benefit, particularly for daily ENDS use, whereas the remainder found no statistically significant association (Xu et al., 2026). Substantial heterogeneity in sample definitions, exposure and outcome measures, covariate selection, and analytic methods limited comparability across studies and precluded consistent conclusions about effectiveness. Importantly, the observational approaches used in these studies did not convincingly establish causal effects, highlighting the need for more rigorous causal methods to determine whether ENDS use facilitates smoking cessation. Evidence is even more limited regarding whether the effects of ENDS use vary across population subgroups. Existing studies, mainly based on earlier-generation ENDS or cross-sectional data, have generally found little evidence of effect modification by age (Jackson et al., 2024; Kosterman et al., 2021), sex (Kosterman et al., 2021; Verplaetse et al., 2019), or race/ethnicity (Kosterman et al., 2021). However, these studies also lacked the rigor necessary to defend a causal interpretation, leaving important uncertainty about whether ENDS facilitate cessation and for whom they may be most effective.

Therefore, we used causal inference methods to estimate the effect of ENDS on subsequent cigarette cessation using rigorous causal methods. Using longitudinal observational data from the PATH Study collected in Waves 4 – 6 (2016 – 2021), we applied Bayesian Additive Regression Trees (BART) to estimate causal effects (Hill, 2011) to address the following questions.

> **Q1** Average Exposure Effect (ATE): Among established cigarette smokers, what would be the average change in the percentage of subsequent cigarette smoking if they used ENDS (a) daily versus not at all (Daily ENDS use vs. No use) or (b) on some days versus not at all (Some-day ENDS use vs. No use)?
>
> **Q2** Group-specific ATE: Are the effects identified in Q1 moderated by participants’ sex, race/ethnicity, annual household income, sexual orientation, internalization, externalization, or age?

## Methods

### Data source and study sample

The PATH Study is one of the most comprehensive initiatives undertaken by the National Institutes of Health (NIH) and the U.S. Food and Drug Administration (FDA) to monitor population-level tobacco use and relevant health outcomes in the context of federal tobacco regulation. Since 2013, the PATH Study has followed a nationally representative sample of youth (aged 12-17 years; n = 13,651) and adults (aged ≥ 18 years; n = 32,320), including both smokers and non-smokers, with follow-up assessments approximately every one to two years. Each wave collects more than 1,000 variables covering tobacco and nicotine use, health outcomes, and a broad range of individual, product, social, and environmental factors corresponding to the classical Host–Agent–Vector–Environment framework. Additional details on the PATH Study design and methods have been reported elsewhere (Hyland et al., 2017).

The present analysis included established current cigarette smokers who participated in Waves 4 (12/2016-01/2018), 5 (12/2018 – 11/2019), and 6 (03/2021 – 11/2021). Eligible participants have smoked >100 cigarettes in their lifetime at Wave 4; reported past-30-day smoking at both Waves 4 and 5; have available ENDS exposure status at Wave 5; and reported current smoking status outcome at Wave 6. After excluding participants with missing baseline covariate data, the final analytical sample consisted of 4792 participants. Sample selection is presented in Figure 1.

**Figure 1.**
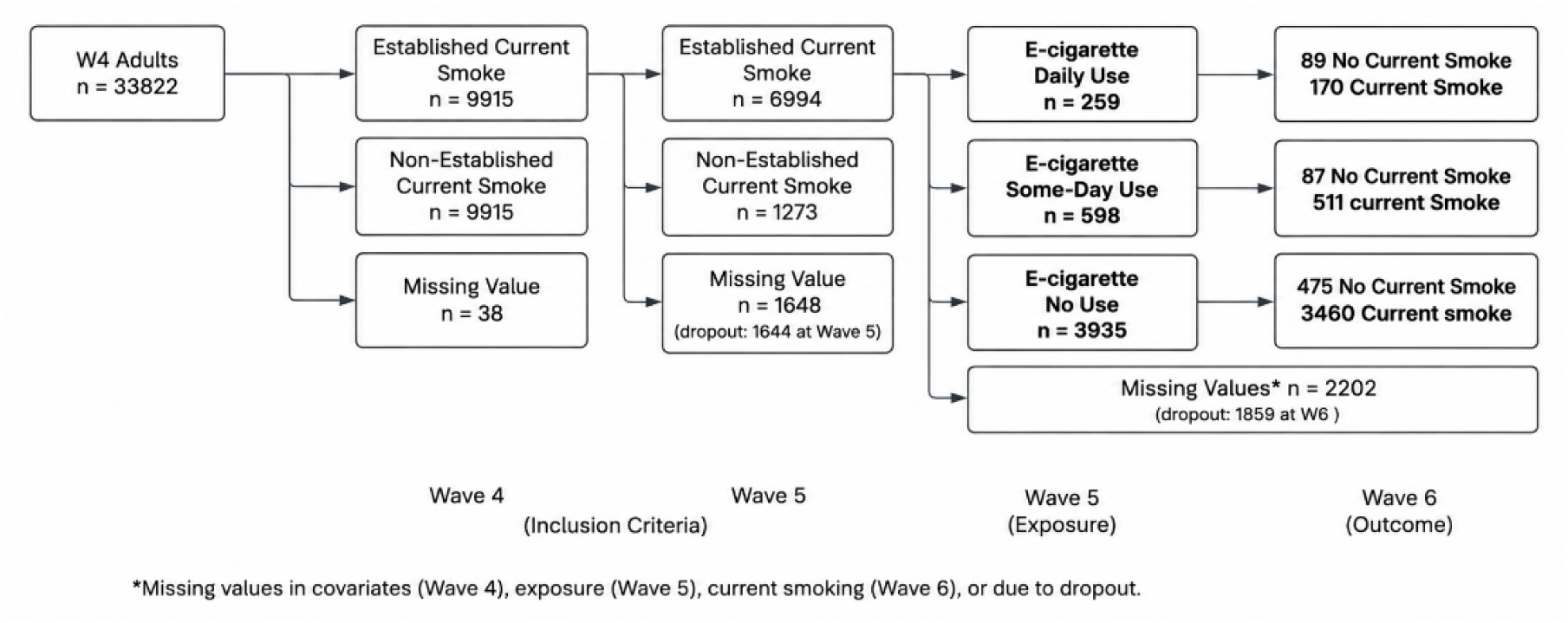
Flowchart of inclusion criteria of study sample. * Missing values in covariates (Wave 4), exposure (Wave 5), current smoking (Wave 6), or due to dropout (at Wave 6). Bold text indicates the analytic study sample.

## Measures

To strengthen the temporal basis for causal inference, we defined a clear sequence in which covariates preceded ENDS exposure, which in turn preceded the smoking outcome. This temporal sequence helps to reduce potential bias from adjusting for post-exposure variables that could function as mediators, colliders, or reflect bidirectional relationships (Rubin, 2008, 2009; Xu et al., 2022). Details of study variables are presented in Table S1.

**ENDS exposure** (Wave 5). Using a PATH-derived variable, participants were first classified by any past-30-day ENDS use. Among participants reporting ENDS use, frequency of use was determined based on the number of days ENDS were used during the past 30 days. ENDS exposure was categorized as no use (0 days), some-day use (1 – 29 days), or daily use (30 days).

**Cigarette smoking outcome** (Wave 6). Using a PATH-derived variable, participants were classified as any past-30-day smoking (assigned a value “1”), indicating no cigarette cessation and no past-30-day smoking (assigned a value “0”), indicating cessation.

**Covariates** (Wave 4). We adjusted for 25 pre-exposure covariates selected based on their potential association with both ENDS use and cigarette smoking (Harlow et al., 2022; Kasza et al., 2021; Xu et al., 2023, 2026). Pre-exposure covariates included sociodemographic (i.e., age group, sex, race/ethnicity, educational attainment, annual household income, region, sexual orientation), tobacco-related perceptions and environmental factors (i.e., absolute harm perception of cigarette smoking, relative harm perception of ENDS use compared to cigarette smoking, household tobacco use policy, family tobacco use, secondhand smoking exposure), tobacco use history and nicotine dependence (i.e., ENDS use, other combustible tobacco use [including cigar, pipe tobacco, and hookah], time to first smoking after waking, nicotine dependence, cigarette quit intention), and health and behavioral covariates (i.e., mental health status [internalization, externalization], body mass index [BMI], chronic disease [heart, respiratory disease, cancer] history, and past month marijuana use and alcohol use).

## Analytic Strategy

Descriptive statistics of study variables were summarized for the overall analytical sample and by ENDS exposure level. As an initial unadjusted analysis, we used a log-binomial regression with an identity link to examine the association between ENDS exposure and cigarette smoking outcome approximately two years later. Effect estimates were expressed as percentage differences to facilitate comparison with the subsequent BART-based causal effect estimates.

**Bayesian Additive Regression Tree** (BART) is a flexible, nonparametric Bayesian approach that models the conditional distribution of an outcome using an ensemble of regression trees (Chipman, George, & McCulloch, 2010). Rather than imposing a prespecified functional form for the relationship between predictors and the outcome, BART combines the fit from many shallow trees, each contributing incrementally to the overall prediction. Regularizing priors constrain individual trees to reduce overfitting while allowing the ensemble to capture nonlinearities and complex interactions. Model estimation is performed using Markov Chain Monte Carlo (MCMC), yielding posterior distributions that quantify uncertainty in effect estimates.

**BART for causal effects**. BART has been increasingly applied to causal inference (Carnegie, Dorie, & Hill, 2019; Dorie et al., 2022; Hill, Perrett, & Dorie, 2023) following recent methodological developments (Hill, 2011; Hill et al., 2020; Dorie & Hill, 2024). Our primary causal BART model to predict the outcome as a function of ENDS exposure, covariates, and estimated propensity scores (estimates obtained from separate BART models). Including the propensity score in the outcome model can improve confounding adjustment and reduce bias in BART-based causal effect estimation (Hahn, Murray, & Carvalho, 2020). For binary current smoking outcomes, BART used a probit link to map the sum-of-trees prediction onto the probability scale. The fitted outcome model was used to generate posterior distributions of individual-level potential outcomes under each exposure condition. For each model, we fit 10 independent MCMC Chains with 1000 burn-in iterations followed by 500 sampling iterations, and specified an ensemble of 75 trees. Convergence was evaluated by examining trace plots across chains.

In the context of a binary exposure (e.g., Daily vs. no ENDS use), the average exposure effects (ATEs) were calculated by averaging the individual-level differences in these predicted outcomes and were reported as percentage-point differences between exposure conditions. For example, a negative ATE for daily versus some-day ENDS use indicates a lower probability of current smoking. Correspondingly, a higher probability of smoking cessation under daily ENDS use.

*Estimands*. We conducted three parallel sets of analyses, each comparing predicted smoking outcomes across two different ENDS use levels. The primary contrasts were daily versus no ENDS use and some-day versus no ENDS use, with daily versus some-day ENDS use as a secondary contrast. For each contrast, we estimated ATE across the overall sample (Q1), and subgroup-specific ATEs within prespecified subgroups (Q2).

*Identification assumptions and sensitivity tests*. Identification of these causal effects relies on three key assumptions (Hill, 2011). First, we assumed conditional exchangeability (also referred to as no *unmeasured confounding* or *ignorability*): formally this requires that ENDS exposure is independent of the potential outcomes. We included an extensive set of covariates based on the literature to increase the plausibility of this assumption (Xu et al., 2026). Nevertheless, because unmeasured confounding cannot be ruled out in an observational study, in sensitivity tests, we conducted sensitivity analyses to assess the robustness of the estimated effects to potential unmeasured confounding (VanderWeele & Ding, 2017). Specifically, we calculated E-values for the risk ratios corresponding to the reduction in current smoking for each pairwise exposure contrast. E-value quantifies the minimum strength of association that an unmeasured confounder would need to have with both ENDS exposure and current smoking, conditional on the measured confounders, to fully explain away the estimated effect. Larger E-values indicate that stronger (e.g., risk ratio > 3 indicating a large effect size; Oliver, May, & Bell, 2017) unmeasured confounding would be required to explain away the observed effect and, therefore, suggest greater robustness to unmeasured confounding. Second, we assumed *common support* (also referred to as positivity or overlap), which requires that all individuals have a nonzero probability of receiving each exposure level conditional on their covariates, allowing for meaningful causal comparisons. We assessed common support using the approach of Hill and Su (2013), which uses posterior uncertainty in estimated potential outcomes to identify observations lacking adequate empirical counterfactual support. Specifically, observations with unusually high posterior uncertainty for their counterfactual outcomes were considered to have insufficient common support and were excluded from the analysis. This procedure is implemented within the BART estimation algorithm, allowing common support to be assessed based on uncertainty in outcome prediction rather than solely on overlap in propensity scores. Third, we assumed *consistency*, such that each participant’s observed outcome under the observed exposure corresponds to that participant’s potential outcome under that exposure, and is not affected by the ENDS exposure of other participants. This assumption was considered tenable given the extensive set of covariates based on the literature and the study design of the PATH study although we note that the definition of “some exposure” does create some vagueness if the interpretation of subsequent causal effects with this contrast.

**Missing data**. We conducted attrition analyses to examine the characteristics of Wave 4 established current smokers who were lost to follow-up at Waves 5 and 6, respectively. Among participants who completed all three waves (Waves 4–6), a small proportion (6%) with missing covariate or exposure data were excluded from the BART analyses.

### Software

BART analysis for causal inference was conducted using the R *bartCause* package (Dorie & Hill, 2020). Sensitivity analyses for unmeasured confounding using E-values were calculated in Stata (Linden et al., 2020).

## Results

Sample characteristics overall and stratified by ENDS exposure level are presented in Table S2. At Baseline, most participants were aged 18-64 years (95.6%), 54.2% were female, and 62.8% were non-Hispanic White. By design, all participants were established current cigarette smokers at both baseline and exposure waves.

At the exposure wave, 259 participants (5.4%) reported daily ENDS use, of whom 89 (34.4%) reported no current cigarette smoking at the outcome wave. Among 598 participants (12.5%) who reported some-day ENDS use, 87 (14.6%) reported no subsequent current smoking. In comparison, 3935 participants (82.1%) reported no current ENDS use; 475 (12.1%) reported no subsequent current smoking. Attrition analysis indicated that, among established current smokers at Wave 4, participants lost to follow-up at Wave 5 were more likely to be younger (p<.001) and male (p<.0001). Similar demographic differences were observed among participants who were lost to follow-up at Wave 6 or excluded because of missing study variables; however, ENDS use frequency did not differ between participants lost to follow-up and those retained (p = .497), and to report more frequent past-30-day ENDS use (p = .014) than those retained.

In the unadjusted log-binomial model, daily ENDS use was associated with a 21.5-percentage-point lower probability of subsequent cigarette smoking compared with no ENDS use (estimate = –21.5%, 95% CI = (–27.0% to –15.9%)). In contrast, the association for some-day ENDS use was smaller and not statistically significant (estimate = –2.8%, 95% CI = (–5.8% to 0.2%)).

**Q1: Average Exposure Effect**. As shown in Figure 2 (and Table S3), are findings suggest that if our causal assumptions hold, daily ENDS reduced the probability of subsequent current smoking compared with no ENDS use (Daily vs. No use: estimate = –15.7%, 95% CI: – 21.5% to –9.9%) or some-day ENDS use (Daily vs. Some-day use: estimate = –15.0%, 95% CI: – 22.3% to –7.8%). In contrast, the estimated difference between some-day and no ENDS use was smaller (Some-day vs. No use estimate = −0.6%, 95% CI: −3.2% to –2.0%). Across all contrasts, only four participants were excluded because of insufficient common support. Sensitivity analyses indicated that, beyond the measured covariates, an unmeasured pre-exposure covariate would need to be associated with both daily ENDS use and subsequent smoking by risk ratios of at least 3.98 for Daily versus no ENDS use and 3.32 for Daily versus Some–day use to fully explain away the observed effects (Figure S1).

**Figure 2.**
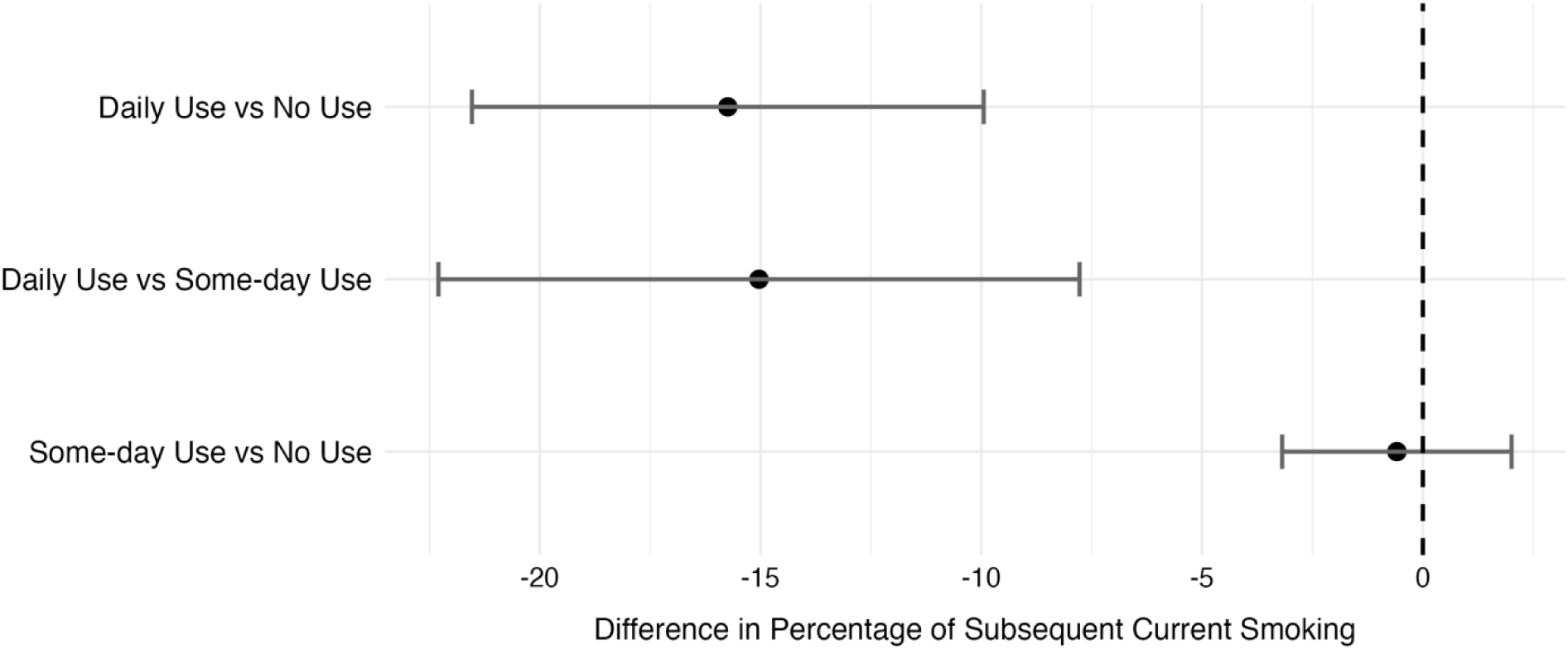
Causal Effects of ENDS Use on Subsequent Current Smoking: Estimates of ATE. *Notes.* ATE = average exposure effect

**Q2: Group-specific ATE.** We further examined whether the estimated effects varied across prespecified subgroups. As shown in Figure 3 (and Table S3), there was no evidence of meaningful effect differences by sex, race/ethnicity, annual household income, sexual orientation, internalization, externalization, or age. Subgroup-specific estimates showed substantial overlap in their 95% credible intervals, suggesting generally consistent ENDS exposure effects across subgroups.

**Figure 3.**
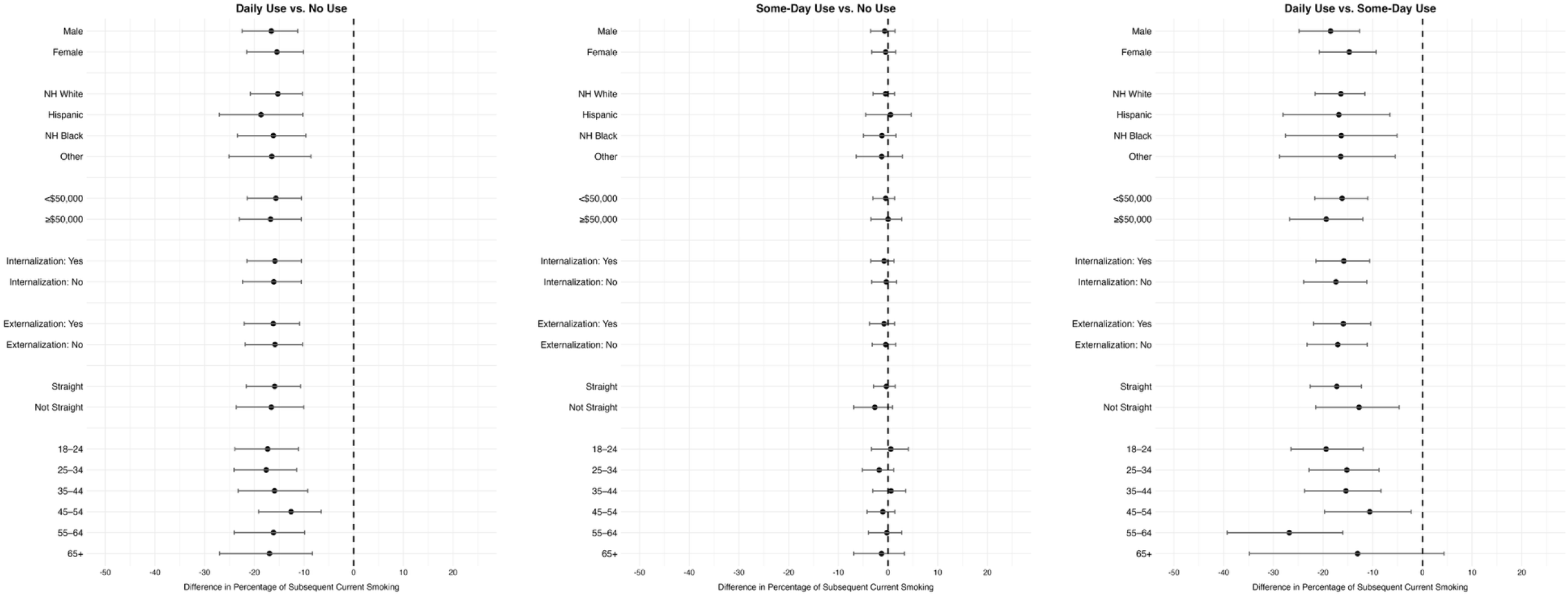
Causal Effects ENDS Use on Subsequent Current Smoking: Estimates of Group-Specific ATE. *Notes.* ATE = average exposure effect.

## Discussion

This study is among the first to apply a causal machine learning approach BART for causal, to estimate the effect of ENDS use on subsequent current smoking status while addressing high-dimensional confounding. Using longitudinal PATH Study data, we focused on a large and diverse sample of established adult smokers who consistently reported past-30-day smoking. To strengthen causal inference, we included an extensive set of 25 pre-exposure covariates selected based on literature and empirical evidence, with all variables defined to preserve the temporal ordering of covariates, exposure, and outcome. BART further reduced reliance on parametric assumptions by flexibly accommodating high-dimensional covariates, nonlinear relationships, and complex interactions. Collectively, these design and analytic features strengthen the basis for causal interpretation of the estimated ENDS exposure effects for the overall sample and are consistent across various subgroups. Although residual confounding cannot be entirely ruled out in an observational study, sensitivity tests suggested that the estimated effects of daily ENDS use were robust to moderate-to-large unmeasured confounding beyond the covariates accounted for in this study. The findings may therefore provide useful evidence for informing public health policy and interventions related to ENDS use and cigarette smoking.

Our findings indicate that, over an approximately two-year period, daily ENDS use decreased subsequent cigarette current use, suggesting that daily ENDS may facilitate smoking cessation. The estimated effect was attenuated relative to the unadjusted estimate from the log-binominal model, suggesting that confounding may have inflated the crude association and underscoring the importance of rigorous confounding adjustment. Some-day ENDS use showed an effect in the same direction in both BART and log-binomial analyses, but of substantially smaller magnitude, with little difference from no ENDS use.

Our study adds evidence on the effects of daily and some-day ENDS on smoking cessation to a literature that remains limited and inconsistent. A review of 18 cohort studies involving 39,147 participants found no overall association between ENDS use and smoking cessation; however, the synthesis did not distinguish between daily and some-day ENDS use. Among the three studies using more recent PATH Study data from Wave 4 onward (Erinoso, et al., 2024; Quach et al., 2025; Sharma et al., 2024), our findings are consistent with Erinoso et al. (2024), who found that daily ENDS use, compared to no use, was associated with greater cigarette abstinence over two years among baseline established adult smokers. In contrast, Sharma et al. found no significant difference in cessation between daily and some-day ENDS users over two years, while Quach et al. (2025) found no association between daily ENDS use and cigarette cessation over four years. These discrepancies may reflect differences in follow-up duration, exposure definitions, analytical approaches, and confounding adjustment.

The generalizability of our findings should be considered in the context of the rapidly evolving ENDS product landscape. The products used during the study period differed substantially from both earlier-generation products (e.g., ciglikes), which generally delivered nicotine less efficiently, and new nicotine-salt pod and disposable devices that became increasingly prominent thereafter (Benowitz et al., 2021; Lindson et al., 2026). At the time of exposure assessment (12/2018 – 11/2019), the US market was dominated by pod-based ENDS products (e.g., JUUL), representing a critical shift in vaping technology and use patterns (Jiang et al., 2023). Newer-generation devices can deliver nicotine more rapidly and efficiently than earlier products, which may influence their effectiveness for cigarette cessation. Accordingly, the observed effects may not generalize uniformly across ENDS product generations. Future studies should directly compare the effectiveness of different ENDS device types and generations for promoting cigarette cessation.

Although daily ENDS use reduced subsequent cigarette smoking, this finding alone does not establish a reduction in tobacco-related harm. Biomarker studies indicate that exclusive ENDS users generally have substantially lower exposure to many tobacco-related toxicants than exclusive cigarette smokers, whereas dual users typically have higher toxicant exposure than exclusive ENDS users (Hartmann-Boyce et al., 2023). Thus, the potential harm-reduction benefit of ENDS depends partly on whether smokers transition completely from cigarettes rather than sustain dual use. ENDS use may also prolong nicotine dependence among individuals who otherwise might achieve complete nicotine abstinence (Auer et al., 2024). Future studies should therefore examine when smokers initiate daily ENDS use, the duration of dual use, and whether daily use facilitates timely and sustained switching away from combustible cigarettes.

Findings on the heterogeneity in the effectiveness of ENDSs for smoking cessation across subgroups should also be interpreted with caution. While the overall study sample is large, some stratified subgroups (particularly daily ENDS users who successfully quit) contain relatively few individuals. This limited number of quitters reduces the information available within these strata, possibly limiting the precision, reliability, and generalizability of the subgroup-specific estimates.

This study has several limitations. First, the observational survey data relied on self-reported measures and may therefore be subject to reporting bias. Second, the validity of the conclusion rests on the assumption that there are no unmeasured confounders. Although our list of pre-treatment covariates was based on a review of the literature, it is likely that some critical factors were ignored. Third, ENDS exposure was treated as a categorical variable measured at a single time point; we did not account for dosage or cumulative use over time. Fourth, we did not account for sampling weights in the BART causal inference analyses of binary outcomes because of current methodological and software limitations. Lastly, we used the most common definition of cigarette cessation (i.e., current non-smoking), without testing alternative definitions. The results may be sensitive to different operationalizations of the outcome or study design.

## Conclusion

Daily ENDS use among established cigarette smokers is associated with subsequent cigarette smoking cessation, compared with some-day or no use. These findings highlight the potential of ENDS as a harm reduction tool for adult smokers. However, further research is needed to better understand the behavioral patterns and long-term health consequences associated with daily ENDS use.

## Data Availability

This study used the Public Use data from the Population Assessment of Tobacco and Health Study. The data (ICPSR 36498) are publicly available at the Inter-university Consortium for Political and Social Research. https://doi.org/10.3886/ICPSR36498.v19.

## Declarations

### Ethics Approval

No IRB involved.

### Informed consent

Not applicable.

### Clinical Trial Registration

Not applicable.

### Author Contributions

SX, JH, YF, and RSN contributed to conceptualization, methodology, and supervision. SX, LC curated the data. SX, JH, LC, and PH contributed to formal analysis, investigation, software, and validation. SX secured funding and managed project administration. SX and RSN provided resources. SX, LC and PH prepared the visualizations. All authors contributed to writing the original draft, and SX, JH, YF, NJ, and RSN contributed to manuscript review and editing.

## Supporting information

Supplementary all

