## Supplementary all for "Do Electronic Nicotine Delivery Systems (ENDS) Help Smokers Quit? A Causal Bayesian Additive Regression Tree Analysis of PATH Study Waves 4 to 6"

Supplementary Materials

Table S1 Description of Study Variables……………………………………………………………………………………….. 2
Table S2 Descriptive Statistics of The Overall Study Sample and by Exposure Group ………………………………………..…….8
Table S3 Causal Effects ENDS Use on Subsequent Smoking: Estimates of ATE and Group-Specific ATE…………………………..10
Figure S1 E-Values Sensitivity Tests for Unmeasured Confounding…………………………...…………………………………11

Table S1 Description of Study Variables

| | PATH Variable | Label | Response Category | Recoded Response Category |
|---|---|---|---|---|
| | | **Covariates (Wave 4)** | | |
| Age Group | R04R_A_AGECAT6 | Age (in years) at the time of interview. | 1 = 18 to 24 years old;<br>2 = 25 to 34 years old;<br>3 = 35 to 44 years old;<br>4 = 45 to 54 years old;<br>5 = 55 to 64 years old;<br>6 = 65 years old or older | 1 = 18 to 24 years old;<br>2 = 25 to 34 years old;<br>3 = 35 to 44 years old;<br>4 = 45 to 54 years old;<br>5 = 55 to 64 years old;<br>6 = 65 years old or older |
| Sex | R04R_A_SEX_IMP | PATH imputed gender | 1 = Male;<br>2 = Female | 0 = Female;<br>1 = Male |
| Race/Ethnicity | R04R_A_RACECAT3 | Race (3 levels) | 1 = White alone;<br>2 = Black alone;<br>3 = Other | (recoded into four binary variables)<br>Hispanic (0 = No; 1= Yes);<br>NH White (0 = No; 1= Yes);<br>NH Black (0 = No; 1= Yes);<br>Other (0 = No; 1= Yes); |
| | R04R_A_HISP_IMP | PATH imputed Hispanic origin | 1 = Hispanic;<br>2 = Not Hispanic | |
| Education Attainment | R04R_A_AM0018_V2 | What is the highest grade or level of school you completed? (5 levels) | 1 = Less than High School<br>2 = GED<br>3 = High school graduate<br>4 = Some college (no degree) or associates degree<br>5 = Bachelor's degree or advanced degree | 1 = Less than high school<br>2 = GED<br>3 = High school graduate<br>4 = Some college (no degree) or associates degree<br>5 = Bachelor's degree or advanced degree |
| Annual Household Income | R04R_A_AM0030 | Which of the following categories best describes your total household income in the past 12 months? | 1 = Less than $10,000<br>2 = $10,000 to $24,999<br>3 = $25,000 to $49,999<br>4 = $50,000 to $99,999<br>5 = $100,000 or more | 0 = $50,000 or more;<br>1 = less than $10,000 - $49,999;<br>2 = missing values |
| | R04_AM0031 | Is your total household income during the past 12 months above or below $50,000? | 1 = Above $50,000<br>2 = Below $50,000 | |
| Region | R04X_CB_REGION | Groupings of states and the District of Columbia that subdivide the United States for the presentation of census data. | 1 = Northeast<br>2 = Midwest<br>3 = South<br>4 = Wes | 1 = Northeast<br>2 = Midwest<br>3 = South<br>4 = West |
| Sex Orientation | R04R_A_SEXORIENT2 | 2 category recode of sexual orientation. | 1 = Lesbian, Gay, Bisexual, Something else;<br>2 = Straight | 0 = Lesbian, Gay, Bisexual, Something else;<br>1 = Straight |

| | | | | |
|---|---|---|---|---|
| Absolute Harm Perception of Cigarette Smoking | R04_AC9050 | How harmful do you think cigarettes are to health? | 1 = Not at all harmful<br>2 = Slightly harmful<br>3 = Somewhat harmful<br>4 = Very harmful<br>5 = Extremely harmful | 1 = Not at all harmful<br>2 = Slightly harmful<br>3 = Somewhat harmful<br>4 = Very harmful<br>5 = Extremely harmful |
| Relative Harm Perception of ENDS Use Compared with Cigarette Smoking | R04_AE1099 | Is using e-cigarettes or other electronic nicotine products less harmful, about the same, or more harmful than smoking cigarettes? | 1 = Less harmful<br>2 = About the same<br>3 = More harmful | 1 = Less harmful<br>2 = About the same<br>3 = More harmful |
| Household Tobacco Use Policy | R04_AR1045 | For tobacco products that are burned, such as cigarettes, cigars, pipes or hookah, which statement best describes the rules about smoking a tobacco product inside your home? | 1 = It is not allowed anywhere or at any time inside my home<br>2 = It is allowed in some places or at some times inside my home<br>3 = It is allowed anywhere and at any time inside my home | 1 = It is not allowed anywhere or at any time inside my home<br>2 = It is allowed in some places or at some times inside my home<br>3 = It is allowed anywhere and at any time inside my home |
| Family Tobacco Use | R04R_A_AX0066 | Does anyone who lives with you now do any of the following: Smoke cigarettes, cigars, cigarillos or filtered cigars, pipe tobacco, use e-products exclusively, use other tobacco products, including smokeless, snus and hookah, or no one living in the home uses tobacco | 1 = Cigarettes, cigars, cigarillos or filtered cigars and pipe tobacco<br>2 = E-products exclusively<br>3 = Other tobacco products, including smokeless, snus and hookah<br>4 = No one living in the home uses tobacco | 0 = No one living in the home uses tobacco;<br>1= Cigarettes, cigars, cigarillos or filtered cigars/ Smokeless or other tobacco only |
| Internalization | | Last time you had significant problems with: | 1 = Past month,<br>2 = 2 to 12 months ago,<br>3 = Over a year ago,<br>4 = Never | = 0 if has less than 2 of (R04_AX0161, R04_AX0162, R04_AX0163, or R04_AX0164) in Past month or 2-12 months ago<br><br>=1 if had 2 or more of (R04_AX0161, R04_AX0162, R04_AX0163, or R04_AX0164) in Past month or 2-12 months ago; |
| | R04_AX0161 | Feeling very trapped, lonely, sad, blue, depressed or hopeless about the future | | |
| | R04_AX0162 | Sleep trouble - such as bad dreams, sleeping restlessly or falling asleep during the day | | |
| | R04_AX0163 | Feeling very anxious, nervous, tense, scared, panicked or like something bad was going to happen | | |
| | R04_AX0164 | Becoming very distressed and upset when something reminded you of the past | | |
| Externalization | | Last time you did the following two or more times: | 1 = Past month;<br>2 = 2 to 12 months ago; | = 0 if has less than 2 of (R04_AX0165, R04_AX0166, |

| | | | | |
|---|---|---|---|---|
| | R04_AX0165 | Lied or conned to get things you wanted or to avoid having to do something | 3 = Over a year ago;<br>4 = Never | R04_AX0167, R04_AX0168, R04_AX0169, R04_AX0250, or R04_AX0251) in Past month or 2-12 months ago<br><br>=1 if had 2 or more of (R04_AX0165, R04_AX0166, R04_AX0167, R04_AX0168, R04_AX0169, R04_AX0250, or R04_AX0251) in Past month or 2-12 months ago; |
| | R04_AX0166 | Had a hard time paying attention at school, work or home | | |
| | R04_AX0167 | Had a hard time listening to instructions at school, work or home | | |
| | R04_AX0168 | Were a bully or threatened other people | | |
| | R04_AX0169, | Started physical fights with other people | | |
| | R04_AX0250 | Felt restless or the need to run around or climb on things | | |
| | R04_AX0251 | Gave answers before the other person finished asking the question | | |
| Heart Disease (12M) | | In past 12 months, doctor, nurse or other health professional said you had: | 1 = Marked<br>2 = Not Marked | Of (R04_AX0111_12M_01 - R04_AX0111_12M_05) in the past 12 months<br><br>0 = No disease,<br><br>1 = have disease, indicate had at least 1 in past 12 months<br><br>2 = missing values, indicate all missing in the past 12 months |
| | R04_AX0111_12M_01 | High blood pressure | | |
| | R04_AX0111_12M_02 | High cholesterol | | |
| | R04_AX0111_12M_03 | Congestive heart failure | | |
| | R04_AX0111_12M_04 | A stroke | | |
| | R04_AX0111_12M_05 | A heart attack (also called myocardial infarction) or needed bypass surgery | | |
| | R04_AX0111_12M_06 | Some other heart condition | | |
| Respiratory Disease (12M) | | In past 12 months, doctor, nurse or other health professional said you had a lung or respiratory condition: | 1 = Marked<br>2 = Not Marked | Of (R04_AX0119_12M_01 - R04_AX0119_12M_05) in the past 12 months<br><br>0 = No disease,<br><br>1 = have disease, indicate had at least 1 in past 12 months |
| | R04_AX0119_12M_01 | COPD | | |
| | R04_AX0119_12M_02 | Chronic bronchitis | | |
| | R04_AX0119_12M_03 | Emphysema | | |

| | | | | |
|---|---|---|---|---|
| | R04_AX0119_12M_04<br>R04_AX0119_12M_05 | Asthma<br>Some other lung or respiratory condition | | 2 = missing values, indicate all missing in the past 12 months |
| Cancer (12M) | R04_AX0144_12M | In past 12 months, doctor, nurse or other health professional told you that you had cancer | 1 = Yes;<br>2 = No | 0 = No;<br>1 = Yes |
| Body Mass Index | R04R_A_BMI | Body mass index (BMI) | (Numerical) | 0 = [18.5, 25)<br>1 = (0, 18.5)<br>2 = [25, 30)<br>3 = [30, ∞) |
| Alcohol P30D Use | R04_AX0673 | Have you used alcohol within the past 30 days? | 1 = Yes; 2 = No | 0 = No; 1 = Yes |
| Marijuana P30D Use | R04_AX0675 | Have you used marijuana, hash, THC, grass, pot or weed within the past 30 days? | 1 = Yes; 2 = No | 0 = No; 1 = Yes |
| Other Combustible Tobacco Use | R04R_A_EVR_PIPE<br>R04R_A_EVR_HOOK<br>R04R_A_EVR_CIGAR | Wave 4 adult respondents who have ever smoked a pipe.<br>Wave 4 adult respondents who have ever smoked a hookah.<br>Wave 4 adult respondents who have ever smoked any cigar product. | 1 = Yes; 2 = No | 0 = No; 1 = Yes |
| Cigarette Quit Intent | R04_AN0240 | Time-frame when planning to quit smoking/using tobacco product(s) for good | 1 = In the next 7 days<br>2 = In the next 30 days<br>3 = In the next 6 months<br>4 = In the next year<br>5 = More than one year from now | 0 = in the next 7 days - 6 months<br>1 = in the next year or more than one year from now |
| Secondhand Smoking Exposure | R04_AX0068 | During the past seven days, about how many hours were you around others who were smoking [whether or not you were smoking yourself]? Include time in your home, in a car, at work, or outdoors. | (Numerical) | (Numerical) |
| Time to First Smoking after Waking | R04_AC1024_NN<br>R04_AC1024_UN | Time to first cigarette after waking on days smoked - Number (current smokers)<br>Time to first cigarette after waking on days smoked - Unit (current smokers) | (Numerical)<br>1 = Minutes after waking<br>2 = Hours after waking | (Numerical) |
| Nicotine Dependence | | Level of agreement: | 1 = Not true of me at all | Rescale 1–5 to 0–100 scale: |

| | | | | |
|---|---|---|---|---|
| | R04_AN0025<br><br>R04_AN0030<br><br>R04_AN0065<br><br>R04_AN0035<br><br>R04_AN0045<br><br>R04_AN0085<br><br>R04_AN0090<br><br>R04_AN0060<br><br>R04_AN0095<br><br>R04_AN0100<br><br>R04_AN0055<br><br>R04_AN0050 | Frequently crave tobacco product(s)<br><br>Urges keep getting stronger if don't/Still have urges to smoke/use tobacco product(s)<br><br>Can only go a couple of hours without smoking/using tobacco product(s)<br><br>Tobacco product(s) control me<br><br>My tobacco product(s) smoking/use is out of control/My urge to smoke/use tobacco product(s) is out of control<br><br>Would find it really hard to stop smoking/using tobacco product(s)<br><br>Would find it hard to stop smoking/using tobacco product(s) for a week<br><br>Usually want to smoke/use tobacco product(s) right after waking up<br><br>After not smoking/using tobacco product(s) for a while, need to smoke/use tobacco product(s) in order to feel less restless and irritable<br><br>After not smoking/using tobacco product(s) for a while, need to smoke/use tobacco product(s) in order to keep self from experiencing any discomfort<br><br>Finds self reaching for tobacco product(s) without thinking about it<br><br>Frequently smoke/use tobacco product(s) without thinking about it<br><br>Smoking/using tobacco product(s) really helps me feel better if feeling down | 2 = 2 on a scale of 1 to 5<br>3 = 3 on a scale of 1 to 5<br>4 = 4 on a scale of 1 to 5<br>5 = Extremely true of me | 1 → 0<br>2 or 3 → 50<br>4 or 5 → 100;<br><br>Then sum and count non-missing rescaled responses.<br><br>Finally Compute average per wave (scaled sum ÷ non-missing count) |

| | | | | |
|---|---|---|---|---|
| | R04_AN0070 | Smoking/using tobacco product(s) helps me think better | | |
| | R04_AN0075 | Would feel alone without my tobacco product(s) | | |
| | R04_AN0080 | | | |
| E-cigarette Use | R04R_A_P30D_EPRODS | Wave 4 adult past-30-day electronic nicotine product use | 1 = Yes; 2 = No | 0 = 0 day<br>1 = 1 – 29 days<br>2 = 30 days |
| | R04_AV1022 | How many of the past 30 days used an electronic nicotine product? | (numeric) | |
| | | **Exposure (Wave 5)** | | |
| E-cigarette Use | R05R_A_P30D_EPRODS | Wave 5 adult past-30-day electronic nicotine product use | 1 = Yes; 2 = No | 0 = 0 day<br>1 = 1 – 29 days<br>2 = 30 days |
| | R05_AV1022 | How many of the past 30 days used an electronic nicotine product? | (numeric) | |
| | | **Outcome (Wave 6)** | | |
| Cigarette Current Smoking | R06R_A_P30D_CIGS | Wave 6 adult respondents who have smoked a cigarette within the past 30 days. | 1 = Yes; 2 = No | 0 = No; 1= Yes |

Table S2. Baseline characteristics of the whole sample and by ENDS exposure level

| Variable | Overall Sample (n = 4792) | No Use (n = 3935, 82.1%) | Some-Day Use (n = 598, 12.5%) | Daily Use (n = 259, 5.4%) |
|---|---|---|---|---|
| **Age Group** | | | | |
| 18 to 24 years | 626 (13.1%) | 399 (10.1%) | 141 (23.6%) | 86 (33.2%) |
| 25 to 34 years | 1142 (23.8%) | 879 (22.3%) | 183 (30.6%) | 80 (30.9%) |
| 35 to 44 years | 889 (18.6%) | 733 (18.6%) | 111 (18.6%) | 45 (17.4%) |
| 45 to 54 years | 976 (20.4%) | 844 (21.4%) | 104 (17.4%) | 28 (10.8%) |
| 55 to 64 years | 851 (17.8%) | 795 (20.2%) | 40 (6.7%) | 16 (6.2%) |
| 65+ years | 308 (6.4%) | 285 (7.2%) | 19 (3.2%) | 4 (1.5%) |
| **Sex** | | | | |
| Female | 2595 (54.2%) | 2133 (54.2%) | 337 (56.4%) | 125 (48.3%) |
| Male | 2197 (45.8%) | 1802 (45.8%) | 261 (43.6%) | 134 (51.7%) |
| **Race/Ethnicity** | | | | |
| Hispanic | 622 (13.0%) | 515 (13.1%) | 86 (14.4%) | 21 (8.1%) |
| NH White | 3010 (62.8%) | 2431 (61.8%) | 379 (63.4%) | 200 (77.2%) |
| NH Black | 828 (17.3%) | 730 (18.6%) | 80 (13.4%) | 18 (6.9%) |
| Others | 332 (6.9%) | 259 (6.6%) | 53 (8.9%) | 20 (7.7%) |
| **Education Attainment** | | | | |
| Less than high school | 832 (17.4%) | 700 (17.8%) | 92 (15.4%) | 40 (15.4%) |
| GED | 590 (12.3%) | 501 (12.7%) | 67 (11.2%) | 22 (8.5%) |
| High school graduate | 1235 (25.8%) | 1025 (26.0%) | 144 (24.1%) | 66 (25.5%) |
| Some college | 1651 (34.5%) | 1316 (33.4%) | 227 (38.0%) | 108 (41.7%) |
| Bachelor’s or higher degree | 484 (10.1%) | 393 (10.0%) | 68 (11.4%) | 23 (8.9%) |
| **Annual Household Income** | | | | |
| 0 - $50,000 | 1127 (23.5%) | 910 (23.1%) | 141 (23.6%) | 76 (29.3%) |
| >= $50,000 | 3553 (74.1%) | 2934 (74.6%) | 443 (74.1%) | 176 (68.0%) |
| Missing | 112 (2.3%) | 91 (2.3%) | 14 (2.3%) | 7 (2.7%) |
| **Region** | | | | |
| Northeast | 681 (14.2%) | 560 (14.2%) | 82 (13.7%) | 39 (15.1%) |
| Midwest | 1358 (28.3%) | 1105 (28.1%) | 180 (30.1%) | 73 (28.2%) |
| South | 1918 (40.0%) | 1587 (40.3%) | 228 (38.1%) | 103 (39.8%) |
| West | 835 (17.4%) | 683 (17.4%) | 108 (18.1%) | 44 (17%) |
| **Sex-orientation** | | | | |
| Not straight | 480 (10.0%) | 334 (8.5%) | 99 (16.6%) | 47 (18.1%) |
| Straight | 4294 (89.6%) | 3586 (91.1%) | 498 (83.3%) | 210 (81.1%) |
| Missing | 18 (0.4%) | 15 (0.4%) | 1 (0.2%) | 2 (0.8%) |
| **Absolute Harm Perception of Cigarette Smoking** | | | | |
| Not at all harmful | 92 (1.9%) | 67 (1.7%) | 17 (2.8%) | 8 (3.1%) |
| Slightly harmful | 241 (5.0%) | 196 (5%) | 25 (4.2%) | 20 (7.7%) |
| Somewhat harmful | 1089 (22.7%) | 904 (23%) | 142 (23.7%) | 43 (16.6%) |
| Very harmful | 1746 (36.4%) | 1425 (36.2%) | 224 (37.5%) | 97 (37.5%) |
| Extremely harmful | 1624 (33.9%) | 1343 (34.1%) | 190 (31.8%) | 91 (35.1%) |
| **Relative Harm Perception of ENDS Use Compared with Cigarette Smoking** | | | | |
| Less harmful | 1044 (21.8%) | 756 (19.2%) | 173 (28.9%) | 115 (44.4%) |
| About the same | 3242 (67.7%) | 2738 (69.6%) | 377 (63%) | 127 (49.0%) |
| More harmful | 423 (8.8%) | 368 (9.4%) | 41 (6.9%) | 14 (5.4%) |
| Missing | 83 (1.7%) | 73 (1.9%) | 7 (1.2%) | 3 (1.2%) |
| **House Tobacco Use Policy** | | | | |
| Not allowed anywhere/anytime | 2510 (52.4%) | 2034 (51.7%) | 313 (52.3%) | 163 (62.9%) |
| Allowed in some places or at sometimes | 1201 (25.1%) | 998 (25.4%) | 156 (26.1%) | 47 (18.1%) |
| Allowed in anywhere/at any time | 1081 (22.6%) | 903 (22.9%) | 129 (21.6%) | 49 (18.9%) |
| **Family Tobacco Use** | | | | |

| Variable | | Overall Sample (n = 4792) | No Use (n = 3935, 82.1%) | Some-Day Use (n = 598, 12.5%) | Daily Use (n = 259, 5.4%) |
|---|---|---|---|---|---|
| | No | 1480 (30.9%) | 1225 (31.1%) | 177 (29.6%) | 78 (30.1%) |
| | Yes | 2492 (52.0%) | 1998 (50.8%) | 348 (58.2%) | 146 (56.4%) |
| | Missing | 820 (17.1%) | 712 (18.1%) | 73 (12.2%) | 35 (13.5%) |
| **Internalization** | | | | | |
| | No | 2336 (48.7%) | 1998 (50.8%) | 247 (41.3%) | 91 (35.1%) |
| | Yes | 2450 (51.1%) | 1932 (49.1%) | 350 (58.5%) | 168 (64.9%) |
| | Missing | 6 (0.1%) | 5 (0.1%) | 1 (0.2%) | 0 (0%) |
| **Externalization** | | | | | |
| | No | 2898 (60.5%) | 2493 (63.4%) | 296 (49.5%) | 109 (42.1%) |
| | Yes | 1889 (39.4%) | 1437 (36.5%) | 302 (50.5%) | 150 (57.9%) |
| | Missing | 5 (0.1%) | 5 (0.1%) | 0 (0%) | 0 (0%) |
| **Heart Disease in P12M**[a] | | | | | |
| | No | 1642 (34.3%) | 1320 (33.5%) | 230 (38.5%) | 92 (35.5%) |
| | Yes | 1097 (22.9%) | 957 (24.3%) | 101 (16.9%) | 39 (15.1%) |
| | Missing | 2053 (42.8%) | 1658 (42.1%) | 267 (44.6%) | 128 (49.4%) |
| **Respiratory Disease in P12M**[a] | | | | | |
| | No | 2132 (44.5%) | 1781 (45.3%) | 250 (41.8%) | 101 (39.0%) |
| | Yes | 608 (12.7%) | 497 (12.6%) | 81 (13.5%) | 30 (11.6%) |
| | Missing | 2052 (42.8%) | 1657 (42.1%) | 267 (44.6%) | 128 (49.4%) |
| **Cancer in P12M**[a] | | | | | |
| | No | 2679 (55.9%) | 2228 (56.6%) | 323 (54.0%) | 128 (49.4%) |
| | Yes | 62 (1.3%) | 53 (1.3%) | 7 (1.2%) | 2 (0.8%) |
| | Missing | 2051 (42.8%) | 1654 (42.0%) | 268 (44.8%) | 129 (49.8%) |
| **BMI** | | | | | |
| | Normal Weight | 1553 (32.4%) | 1242 (31.6%) | 216 (36.1%) | 95 (36.7%) |
| | Underweight | 113 (2.4%) | 96 (2.4%) | 11 (1.8%) | 6 (2.3%) |
| | Overweight | 1494 (31.2%) | 1251 (31.8%) | 172 (28.8%) | 71 (27.4%) |
| | Obesity | 1632 (34.1%) | 1346 (34.2%) | 199 (33.3%) | 87 (33.6%) |
| **Alcohol Use in P30D**[b] | | | | | |
| | No | 1955 (40.8%) | 1633 (41.5%) | 217 (36.3%) | 105 (40.5%) |
| | Yes | 2757 (57.5%) | 2240 (56.9%) | 368 (61.5%) | 149 (57.5%) |
| | Missing | 80 (1.7%) | 62 (1.6%) | 13 (2.2%) | 5 (1.9%) |
| **Marijuana Use in P30D**[b] | | | | | |
| | No | 3133 (65.4%) | 2675 (68.0%) | 309 (51.7%) | 149 (57.5%) |
| | Yes | 1458 (30.4%) | 1086 (27.6%) | 268 (44.8%) | 104 (40.2%) |
| | Missing | 201 (4.2%) | 174 (4.4%) | 21 (3.5%) | 6 (2.3%) |
| **Other Combustible Tobacco Use** | | | | | |
| | No | 1056 (22.0%) | 966 (24.5%) | 60 (10.0%) | 30 (11.6%) |
| | Yes | 2636 (55.0%) | 2183 (55.5%) | 317 (53.0%) | 136 (52.5%) |
| | Missing | 1100 (23.0%) | 786 (20.0%) | 221 (37.0%) | 93 (35.9%) |
| **Cigarette Quit Intention** | | | | | |
| | No | 2473 (51.6%) | 2025 (51.5%) | 315 (52.7%) | 133 (51.4%) |
| | Yes | 1230 (25.7%) | 1008 (25.6%) | 143 (23.9%) | 79 (30.5%) |
| | Missing | 1089 (22.7%) | 902 (22.9%) | 140 (23.4%) | 47 (18.1%) |
| **Second-hand Smoking Exposure in P7D**[c] **(in hours)** | | | | | |
| | Mean (SD) | 16.97 (27.85) | 16.86 (27.97) | 18.02 (28.62) | 16.2 (23.95) |
| **Time to First Smoking after Waking (minutes)** | | | | | |
| | Mean (SD) | 79.82 (150.2) | 77.54 (148.77) | 92.8 (159.92) | 84.56 (147.63) |
| **Nicotine dependence** | | | | | |
| | Mean (SD) | 56.01 (28.15) | 56.39 (28.06) | 54.66 (28.04) | 53.4 (29.62) |

Notes. Notes. ENDS Exposure were measured at Wave 5. [a] P12M = past 12 months, [b] P30D = past 30 days, [c] P7D = past 7 days

Table S3 Causal Effects ENDS Use on Subsequent Current Smoking: Estimates of ATE and Group-Specific ATE

| | ENDS Daily Use vs. No Use | | | ENDS Some-Day Use vs. No Use | | | ENDS Daily Use vs. Some-Day Use | | |
|---|---|---|---|---|---|---|---|---|---|
| | estimate | sd | 95% CI | estimate | sd | 95% CI | estimate | sd | 95% CI |
| **ATE (Overall)** | -15.7 | 3.0 | (-21.5 -9.9) | -0.6 | 1.3 | (-3.2 -2.0) | -15.0 | 3.7 | (-22.3 -7.8) |
| **Group-Specific ATE** | | | | | | | | | |
| **Sex** | | | | | | | | | |
| Male | -16.7 | 2.9 | (-22.7 -11.2) | -0.9 | 1.3 | (-3.6 1.4) | -18.4 | 3.0 | (-24.3 -12.7) |
| Female | -15.5 | 3.0 | (-21.5 -10.0) | -0.6 | 1.2 | (-3.3 1.5) | -14.8 | 2.9 | (-20.6 -9.1) |
| **Race/Ethnicity** | | | | | | | | | |
| NH White | -15.4 | 2.7 | (-21.0 -10.4) | -0.7 | 1.1 | (-3.1 1.3) | -16.4 | 2.6 | (-21.8 -11.4) |
| Hispanic | -18.8 | 4.4 | (-27.8 -10.3) | 0.2 | 2.3 | (-4.5 4.5) | -17.2 | 5.5 | (-28.0 -6.5) |
| NH Black | -16.2 | 3.6 | (-23.7 -9.5) | -1.5 | 1.7 | (-5.1 1.6) | -15.8 | 5.6 | (-27.6 -5.1) |
| Other | -16.7 | 4.2 | (-25.1 -8.6) | -1.5 | 2.3 | (-6.1 2.9) | -16.6 | 5.9 | (-28.8 -5.5) |
| **Annual Household Income** | | | | | | | | | |
| <$50,000 | -15.8 | 2.8 | (-21.5 -10.6) | -0.7 | 1.1 | (-3.1 1.2) | -16.1 | 2.7 | (-21.3 -10.8) |
| >=$50,000 | -16.8 | 3.3 | (-23.5 -10.4) | -0.1 | 1.6 | (-3.5 2.8) | -19.1 | 3.7 | (-26.3 -12.0) |
| **Internalization** | | | | | | | | | |
| Yes | -16.0 | 2.8 | (-21.9 -10.8) | -0.9 | 1.2 | (-3.5 1.1) | -15.9 | 2.8 | (-21.2 -10.4) |
| No | -16.2 | 3.0 | (-22.4 -10.6) | -0.6 | 1.3 | (-3.4 1.6) | -17.3 | 3.2 | (-23.9 -11.2) |
| **Externalization** | | | | | | | | | |
| Yes | -16.4 | 2.9 | (-22.3 -10.9) | -1.0 | 1.3 | (-3.6 1.3) | -16.0 | 2.9 | (-21.7 -10.4) |
| No | -15.9 | 3.0 | (-21.9 -10.4) | -0.6 | 1.2 | (-3.3 1.5) | -17.0 | 3.0 | (-23.2 -11.4) |
| **Sexual Orientation** | | | | | | | | | |
| Straight | -16.0 | 2.8 | (-21.8 -10.7) | -0.5 | 1.1 | (-3.0 1.4) | -17.2 | 2.7 | (-22.5 -12.1) |
| Not Straight | -16.8 | 3.6 | (-24.1 -9.8) | -2.7 | 2.0 | (-6.9 1.1) | -12.9 | 4.3 | (-21.5 -4.7) |
| **Age Group (in years)** | | | | | | | | | |
| 18 - 24 | -17.7 | 3.4 | (-24.3 -11.1) | 0.4 | 1.9 | (-3.3 3.9) | -19.2 | 3.7 | (-26.4 -12.3) |
| 25 - 34 | -17.7 | 3.3 | (-24.5 -11.4) | -2.0 | 1.6 | (-5.3 1.0) | -15.4 | 3.7 | (-22.4 -8.0) |
| 35 - 44 | -16.3 | 3.6 | (-23.3 -9.3) | 0.4 | 1.7 | (-3.2 3.6) | -15.5 | 4.0 | (-23.7 -7.7) |
| 45 -54 | -12.7 | 3.3 | (-19.2 -6.5) | -1.2 | 1.4 | (-4.2 1.4) | -10.9 | 4.5 | (-19.7 -3.0) |
| 55 - 64 | -16.3 | 3.6 | (-23.9 -9.7) | -0.4 | 1.7 | (-4.2 2.5) | -26.8 | 6.2 | (-39.3 -14.3) |
| >= 65 | -17.0 | 4.8 | (-26.6 -8.3) | -1.5 | 2.6 | (-6.9 3.3) | -14.6 | 9.9 | (-34.8 4.3) |

Notes. ATE = average treatment/exposure effect; NH White = Non-Hispanic White; NH Black = Non=Hispanic Black; sd = standard deviation; CI = credible intervals. Estimates represent percentage-point differences in the probability of subsequent current smoking between exposure conditions.

Figure S1 E-values sensitivity tests for unmeasured confounding

<table>
<tr><th>Exposure contrast</th><th>E-value</th></tr>
<tr><td>Daily vs. No ENDS Use</td><td>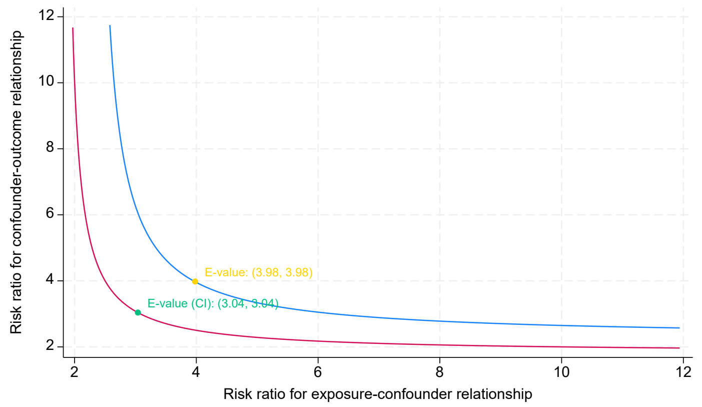
</td></tr>
<tr><td>Some-day vs. No ENDS Use</td><td>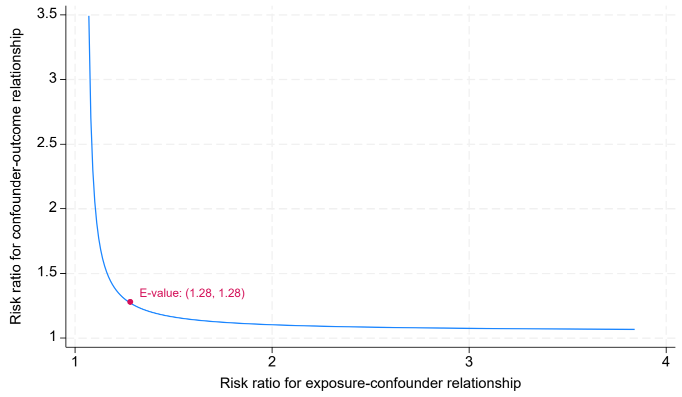
</td></tr>
<tr><td>Daily vs. Some-day ENDS Use</td><td>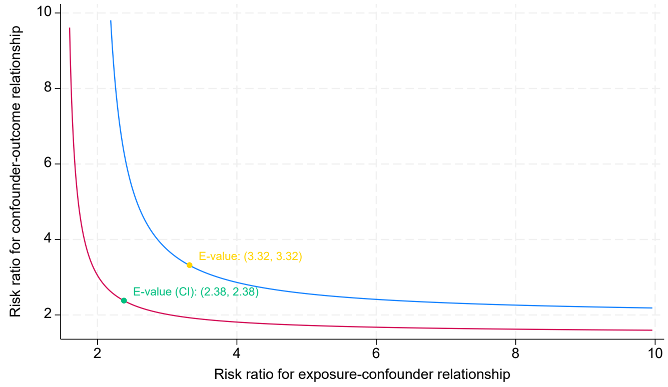
</td></tr>
</table>

Note. Each figure presents an E-value sensitivity analysis for the estimated average exposure effect of ENDS use: daily versus no use, some-day versus no use, and daily versus some-day use, respectively. In the top and bottom panels, E-values of 3.98 and 3.32, respectively, indicate that an unmeasured confounder would need to be associated with both ENDS exposure and the outcome by risk ratios of at least 3.98 and 3.32, respectively, conditional on the measured covariates, to fully explain away the observed effects. These findings suggest that the estimated effects are relatively robust to moderate-to-large unmeasured confounding. In contrast, the middle panel shows a substantially smaller E-value of 1.28 for some-day versus no ENDS use, indicating that relatively weak unmeasured confounding could potentially explain away the observed effect. Thus, the estimated effect of some-day ENDS use was less robust to potential unmeasured confounding. No confidence-interval E-value is shown in the middle panel because the confidence interval for the estimated exposure effect includes the null.
